# Residual immunity and seasonality of an epidemic

**DOI:** 10.1101/2024.05.31.24308217

**Authors:** Siyu Chen, David Sankoff

## Abstract

We present a dynamical model of the onset and severity of cyclical epidemic disease taking account only of seasonal boosts of antibody during the infectious season and residual immunity remaining from one season to the next. We also compile data from public health sources on the annual number of cases of influenza A and peak infectivity month over a quarter century. In these data, we discover that there is a negative correlation between the change in number of cases from one year to the next and the shift of peak infectivity month between the two seasons, although this does not extend to a prediction of epidemic timing or case number based on the the previous season’s statistics. Simulating the mathematical model, we discover that there is also a negative correlation between the change in titer from one season to the next and the shift of peak infectivity month between the two seasons, suggesting that the empirical results can be explained by our minimal boost-and-wane model. In addition, the model predicts that suppressing the epidemic for one season, or witnessing a strong surge for one season, both have lasting effects for a number of successive seasons.

## Introduction

We propose a minimal model of the immunity to a seasonal epidemic disease, exemplified by influenza, taking account only of the exposure-induced boosts during the infectious season and the waning of residual immunity remaining from one season to the next. This offers a direct approach to resolving the contentious issue of how much the timing and severity of one season’s epidemic affects the onset and severity of the next season [1]. Classical models deriving from Kermack–McKendrick [2] focus on the individuals in a population, whether they are susceptible to infection, exposed to the pathogen, symptomatic, hospitalized, in ICU, recovered or deceased. Populations are subdivided into compartments by age, sex, socioeconomic level and/or geographical location. These models calculate the number of individuals in different compartments and account for their movements from one group to another driven by the immunity level (e.g., [3]), concentrating with few exceptions [4] on the details of a single season. Departing from these models, a direct accounting for the population antibody against the implicated pathogen should track the risk trajectory of the epidemic, avoiding the numerous parameters required in compartmental models, and facilitate the study of the time-course of the epidemic across many seasons.

Our model features an exponential waning process during and between annual seasons, imposed on the temporal distribution of antibody boost, reflecting exposures during one season. The latter distribution, especially the timing of peak infectivity, interacting with the waning function, is all that is necessary to reproduce, in mathematically tractable form, the mechanical cycle of immunity characteristic of seasonal infectious disease. We can naturally iterate the cyclical boost-and-wane process to simulate immunity trajectories over many years and thus quantify the relationship between residual immunity and the time elapsed between annual infectivity peaks, namely a relation between end-of-season titer and the length of the interval between annual infectivity peaks.

Determining the onset of epidemic outbreak, whether of an emerging, newly recognized, virulent pathogen [5] or of a recurrent seasonal infection [6, 7, 8], is a vexing problem both for epidemiological research and for population health planning. This has been stressed for a variety of recurrent viral epidemics worldwide [9, 10, 11]. Although the onset and severity of an epidemic are largely unpredictable, there was widespread prediction that the lowering of population immunity against influenza [12] and RSV [13] as a side-effect of non-pharmaceutical interventions during the COVID-19 pandemic (masking, testing, lockdown, isolation,…) would lead to early onset and increased severity of epidemics of these diseases after the pandemic.

To provide an initial assessment of these ideas, we considered a quarter-century of epidemic influenza A statistics from several public health repositories [14, 15, 16, 17]. We compared these statistics and the COVID-19-era predictions with the behaviour of our boost-and-wane model. The result is that the influence of one season’s timing and severity on the next season’s timing and severity is barely perceptible, if at all, in the statistical record, and is not predicted by our model. Moreover in the three post-COVID-19 years, the influenza A seasons in the US and much of the world only returned to levels commensurate with, or less than pre-COVID-19 activity, although the Canada did undergo stronger rebound levels two years later, as did the UK.

Despite the equivocal evidence of the influence of one season on the next, simulations of our model does reveal an unexpected connection between the timing and severity of successive seasons; namely that the change of immune level from one season to the next is substantially and negatively correlated with the span of time between the peak activity of the two consecutive season. This surprising trend is also apparent in the US data and is strongly supported in the Canadian case.

Our model can incorporate a probabilistic determination of peak infectivity, and is thus naturally adaptable to hypotheses about the link between peak month and severity predictions and the pre-season titers. This feature enables a series of experiments to show that after a major disruption, such as a complete suppression or substantial elevation, of one season’s epidemic activity, the re-establishment of pre-existing levels of infectivity is not immediate but requires several years to achieve.

The World Health Organization [14], Health Canada [15], The Centre for Disease Control in the US [16], the UK Health Security Agency [17], among many others, publish weekly reports on influenza strains, including variants of influenza A, dating back to the late 1990s. These data on strains, tests and positivity rates, do not of course include total population antibody levels, but we can use total number of cases detected each week/month during an infectious season as a proxy for these levels, while the peak month can be identified from the visual displays. These data are plotted in Figure 1 for Canada and the US.

**Figure 1:**
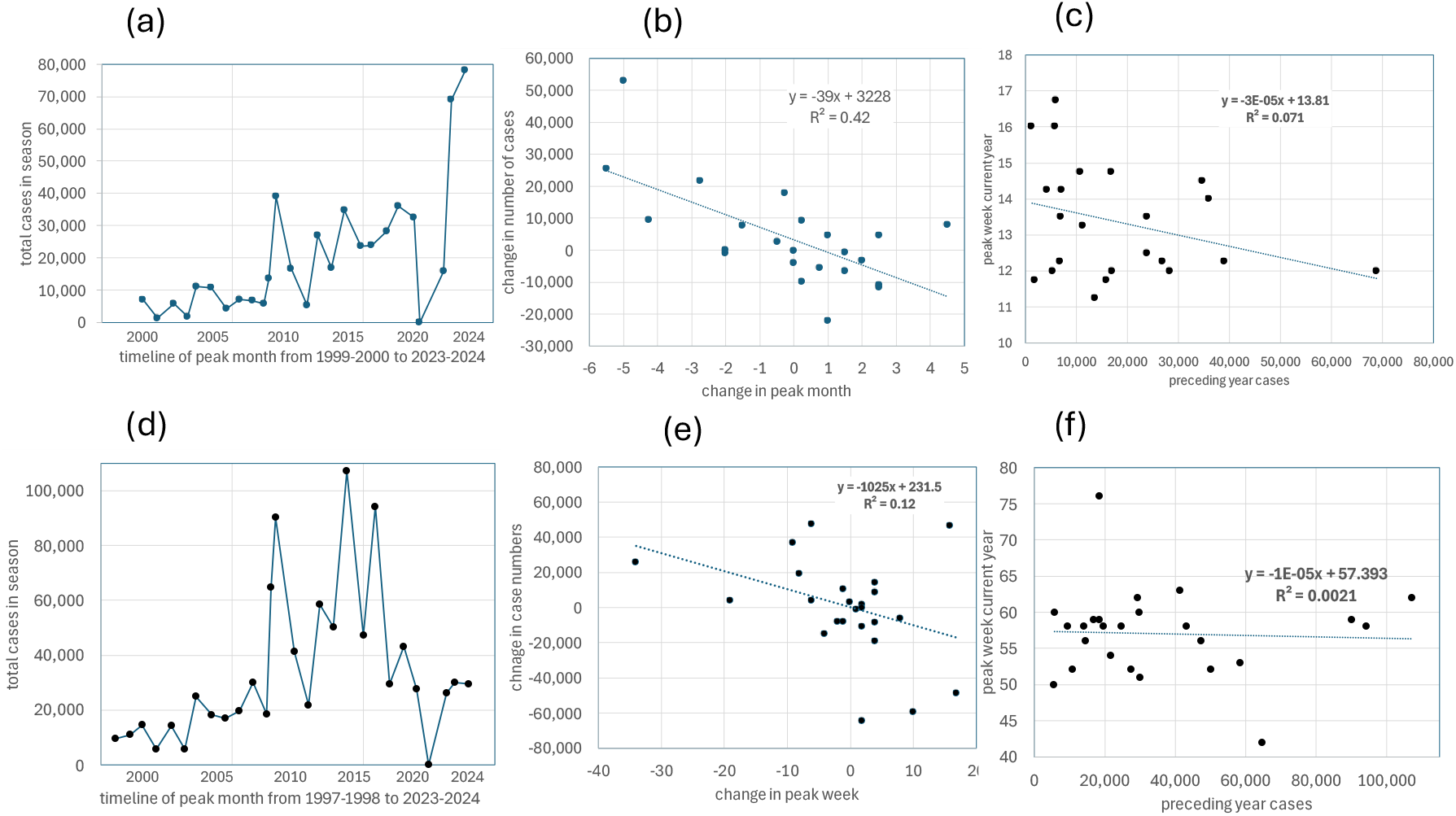
Comparison of influenza epidemic history in Canada (a-c) and the US (d-f). (a) and (d): trajectory of influenza A epidemics over a quarter of a century, x-axis denotes the peak month of the season while y-axis represents the total number of cases in that season. (b) and (e): change in number of cases as a function of change in peak month, and (c) and (f): peak month as a function of number of cases in preceding season.

The timeline of the Canadian and US influenza A experience depicted in Figure 1 shows a clear and dramatic increase in number of cases in the years following the 2009 pandemic. And another major, but not immediate, increase after the COVID-19 pandemic is apparent in the Canadian data and worldwide [14], though not the US. Panels (b) and (e) in Figure 1 show that the change in number of cases from one year to the next is negatively correlated with the shift in peak month, while the panels (c) and (f) does not reveal any strong influence of the peak month of one year on the number of cases in the next year.

## The model

### An infectious cycle for a single season

The average antibody titer of a population will increase because of new exposures but decrease due to antibody waning, as sketched in Figure 2. Our model contains a minimum of elements, namely a probability distribution *f*_*μ*_(*t*) reflecting the stringency of infection, e.g., the rate at which susceptible individuals in a population acquire an infectious disease, and an antibody decay rate *ω*.

**Figure 2:**
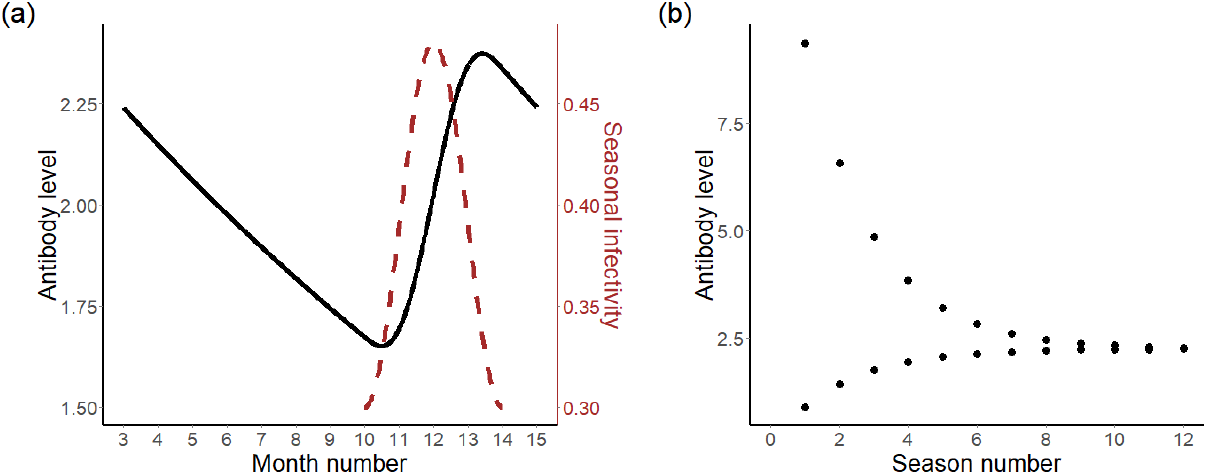
(a) An antibody trajectory over a hypothetical infectious season, March until March the next year, with infectivity curve overlayed. The antibody level starts at 2.24 in March and ends at 2.24 in March of the next year. The dashed brown curve shows the distribution of stringency of infection during the season starting in October ending in February. The peak month of infectivity, predefined at December, corresponds to the steepest increase of antibody level. (b) Antibody trajectories converge to one fixed point at 2.24 over several seasons from two initial levels, 9.37 and 0.88. y-axis is the antibody level at the end of each season, i.e., March of the year.

Then the change of antibody titers between time point *t* = *T*_0_, before an infectious season, and *t* = *T*_1_, after the season, can be calculated by the relation:

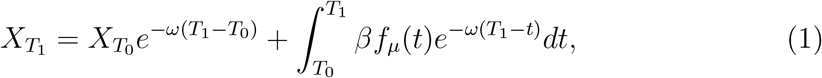

where *β* is the amplitude or severity of the infectious season.

We illustrate with *f*_*μ*_(*t*) as a raised cosine distribution [18], though any distribution *f* with support on an interval within *T*_0_ and *T*_1_, even a Dirac function, would produce results similar to what we will show. The raised cosine function has an amplitude and vertical shift of 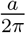, a phase shift of *μ* and a period of 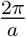, parameterized as:

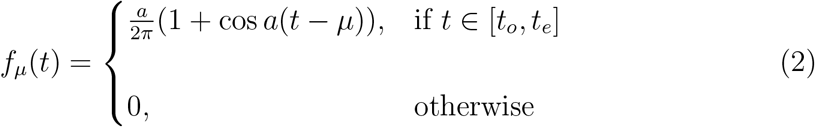

where an infectious season starts from date 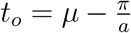 and ends on date 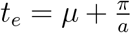, where *μ* is the peak month of the severity of the current infectious season.

From the definition in Equation [2], Equation [1] becomes

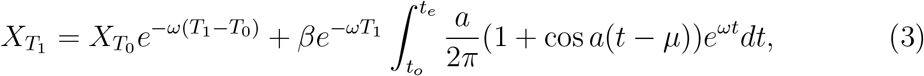

Then, using the definition of *t*_*o*_ and *t*_*e*_, the integral in Equation [3] has closed form

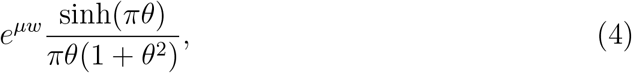

where 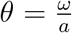. Equation [3] can then be rewritten

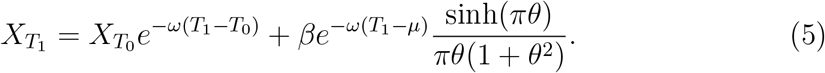

### A fixed point of an iterated model for multiple seasons

Using a given initial titer 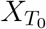 to calculate 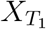 by Equations [1] or [5], and using the calculated 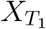 to calculate 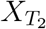, and continuing the same way for 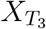, 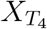, *…*, we have defined a discrete dynamic system, at times *T*_0_ < *T*_1_ < *…*. We can assume *T*_*i*_ − *T*_*i*−1_ > Δ > 0, for some Δ and for all *i* ≥ 1. Given two initial titers 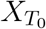 and 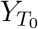, we can see that

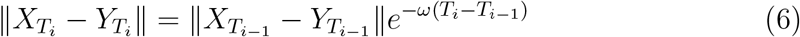

since the integral is constant, for *i* = 1, 2, *…*. Now *e*^−*ω*Δ^ < 1, so this process is contractive, with Lipschitz constant *e*^−*ω*Δ^.

It has a fixed point

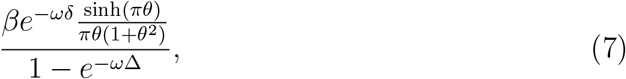

in the simplest case, where *β* = 1, *δ* = *T*_*i*_ − *μ* and Δ = *T*_*i*_ − *T*_*i*−1_ for all *i*.

For example, we may examine the default parameters *β* = 1, 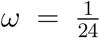, *μ* = 12, 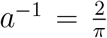, Δ = *T*_*i*_ − *T*_*i*−1_ = 12, *δ* = *T*_*i*_ − *μ* = 3, to determine the fixed point *X* = 2.24. This may be compared to the numerical results in Figure 2(b) tracking two trajectories of the model over 20 seasons.

We note that the fixed point *X* is particularly sensitive to the parameter *μ*, with derivative of form

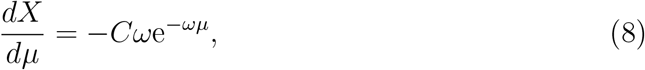

where *C* is a positive constant. This result is important for understanding the tendencies in Figure 1(b) and (e), and the results in Section. Although this result is based on the raised cosine model, similar results can be found for other *f*. In addition, in all the calculations involving fixed points, the factor *β* does not play an important role, since it multiplies *X* in a single year and in the fixed point solution in the same way.

## Long-term trends

### Random peaks

As a benchmark experiment, we concatenated successive instances of the model of a single cycle described in Section to carry out a simulation of 20 recurrent infectious seasons interspersed with quiescent periods for the rest of each cycle (i.e., year). Initialized with a random titer at date *T*_1_ corresponding to the end of a typical infectious period, the peak of next infectious month *m* was chosen from a uniform distribution over a wide range, September (month 9) to April (month 16), and the first iteration was performed at *T*_2_ set to be 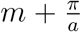, with output 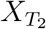. The output 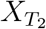 at time *T*_2_ from the first cycle, is then used as the starting titer 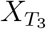 at time *T*_3_ of the second cycle. The peak of next infectious season is again randomly chosen from September to April. This calculation is repeated for the third and subsequent cycles. Figure 3(a) shows five typical trajectories of titers over the 20-year seasons. The randomness in the choice of peaks prevented the process from simply degenerating into a fixed point limit cycle, but maintained a stable pattern of random variation indefinitely.

**Figure 3:**
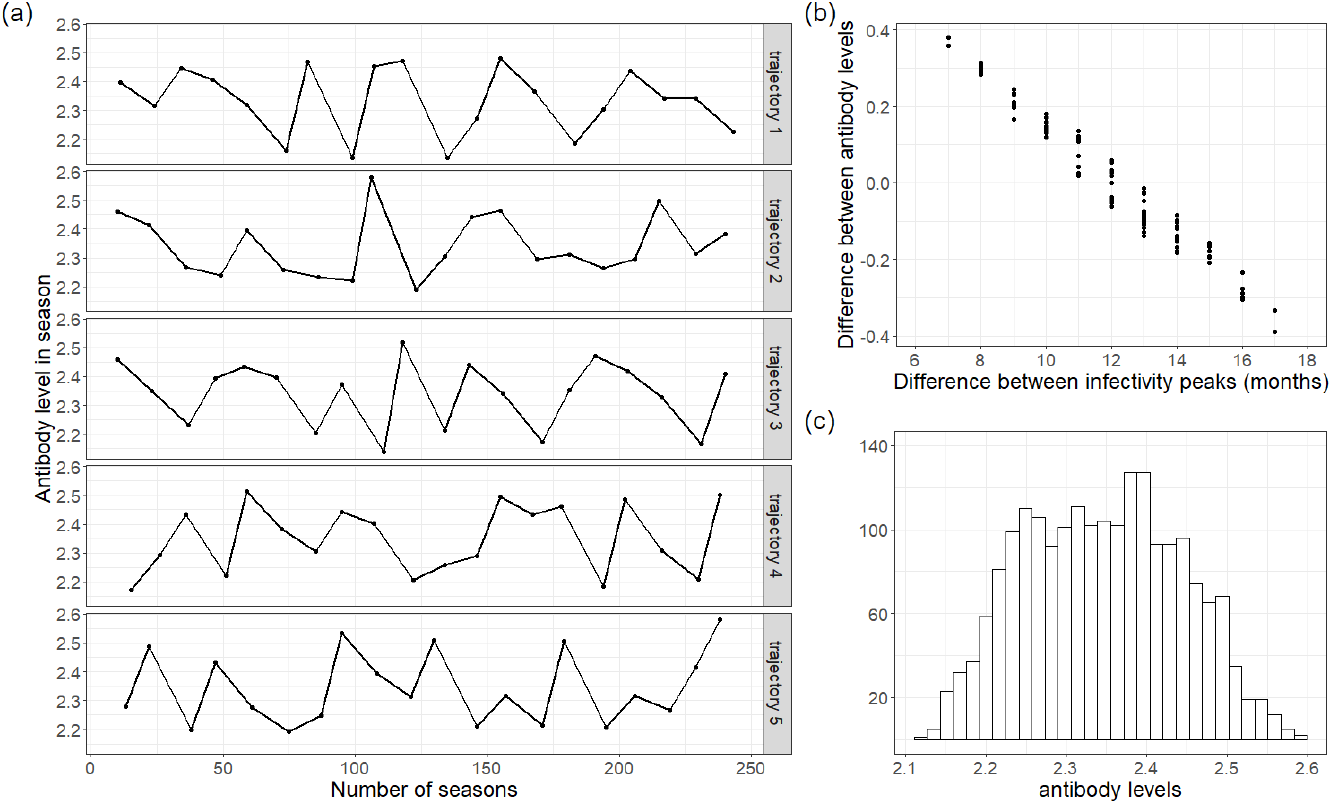
(a) Sample trajectories of 20 seasons produced by iterating the model, five replicates. (b) Association of titer change with length of inter-season times. (c) Long-term distribution of titers of the iterated model.

How can we characterize this pattern formally? There is a contractive model as in Equation [1] corresponding to each value of *μ*. However, as the model is changing with most iterations of the process, the approach to the fixed point(s) is disrupted at each step.

There are *r* different values of *μ*, corresponding to the months from September to April (*r* = 8). If we imposed the unrealistic conditions that the process repeated indefinitely along a fixed trajectory among all the *r* values of *μ* then there would exist a fixed orbit of *r* points, to which the successive titers in the trajectory would converge over time. However, once we allow complete randomness in the successive values of *μ* among the *r* possibilities, as is necessary in our epidemic model, then no such fixed orbit exists. Instead, we can only say that the distribution of titer values converges to a fixed distribution with support [*s*_1_, *s*_*r*_], where *s*_1_ and *s*_*r*_ represent the minimum and maximum fixed points of the *r* models, as in Figure 3(c).

We plotted the change in titer 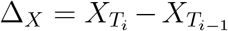 as a function of Δ_*μ*_ = *μ*_*i*_ −*μ*_*i*−1_, the shift in peak infections between the *i* − 1-st and *i*-th cycle. The results in Figure 3(b) show a tight linear relation between the two quantities. The slope is -0.71 titer units/12 months differential, or 0.059 units/month. This compares to log *ω* = 0.042/month. The explanation for this result lies in Equation [8]. The way the titer decreases as a function of *μ* ensures that an early season followed by a late season will result in a low titer, while a late season followed by an early season will have a relatively high titer. Two seasons that occur during the same month will results in an intermediate titer.

### Training for seasonality

The negative correlation between the change of seasonal titer and the time elapsed between successive peaks in Figure 3(b) must be seen as a function only of the waning time between two seasons, since there is no mechanism within the model to affect one year’s peak *μ* as a direct function of the previous year’s output titer 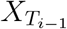. Such a mechanism, however, is widely thought to be of importance to recurrent epidemics, and so we investigate it here.

To train the model as a predictor of seasonality, we made use of the results of the experiment described in Section. Based on each year’s 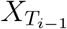, a random choice of next year’s peak month *μ*_*i*_ was effected. This choice involved two steps: The first was the deterministic choice of one of four bins, *B*_1_, *…, B*_4_, with *B*_1_ containing the highest values of 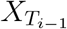, and *B*_4_ containing the lowest values of 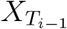. The second step was a uniform random choice among three consecutive months. The months in each bin overlapped those in the adjacent bins, with *B*_1_ containing January through March, *B*_2_ containing December through February, *B*_3_ containing November through January and *B*_4_ containing October through December.

A 200-season experiment could then be initiated by a random choice of 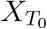. There was no further addition of noise or other intervention over the length of the experiment. Because setting a new peak month determines a different fixed point for 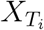, each iteration of the model gives an different output from the previous season’s. Figure 4(a) shows one aspect of the outcome, comparing the titers of each pair of successive seasons. The extent of the scatter is largely the effect of the large overlapping bins from which to choose the peak month.

**Figure 4:**
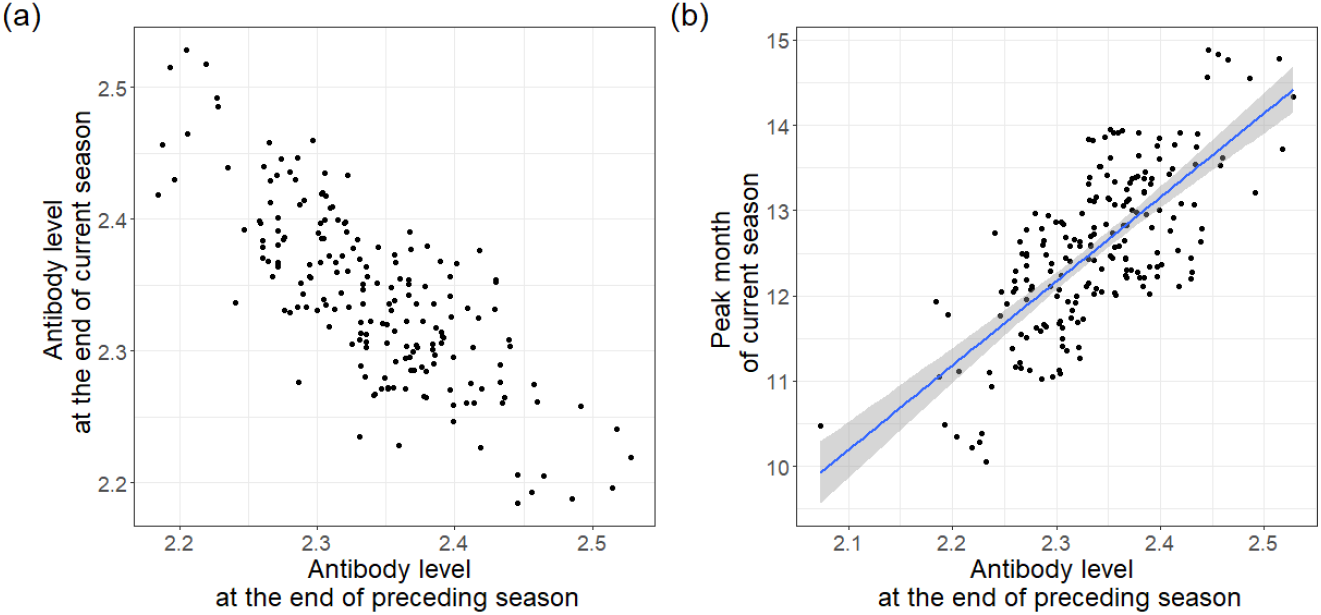
(a) Effect of training on titers of successive seasons. (b) Effect of training on choice of peak month, with regression line.

### Using the model to predict the peak of the next epidemic season

We regress the peak month chosen against the input titer for the data in Section. This produces the following result:

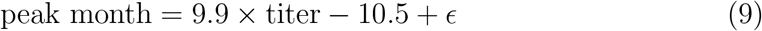

where the normal error *ϵ* has a standard deviation of 0.45. This result can then be used as a predictive tool. Given an end-of-season titer 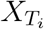, Equation [9] can predict the likely peak month and the shape of the titer distribution during the season. For example, using the parameters in Figure 4(b), an end-of-season titer of 4.7 predicts an early January peak, but distributed with a standard deviation of about two weeks.

### Lasting effects of a missing season

We presented evidence that the dramatically diminished influenza epidemic resulting from the COVID-19 pandemic-inspired non-pharmaceutical interventions in 2020-21 and early 2022 was not followed by an early onset and severe influenza season in 2022-2023, contrary to many predictions [12], based on the putative lack of newly acquired immunity during the pandemic.

From the modeling viewpoint, this aspect of the seasonality-titer relationship, can be expressed by simply introducing a single missing (zero-amplitude) season, and following the trajectory of the process in subsequent years. To accomplish this we started with the model in section above. In one experiment over fifty years, we introduced an ‘*β* = 0’ year every eighth year. In a second experiment we introduced an ‘*β* = 0’ year every four years. We then compared the trajectory from these two experiments with that from the original, unmodified, experiment, where *β* = 1 for every season.

The results show that for the experiment with an 8-year cycle, the effect of a reduced titer, colored orange in Figure 5(a), persisted over five to seven years, before catching up to the sequence of unmodified seasons. On the other hand, with a four-year cycle, colored purple in Figure 5(a), the output titer never quite caught up. Figure 5(b) shows how the titers of the two missing-seasons trajectories gradually increase towards the unmodified (no missing seasons) pattern.

**Figure 5:**
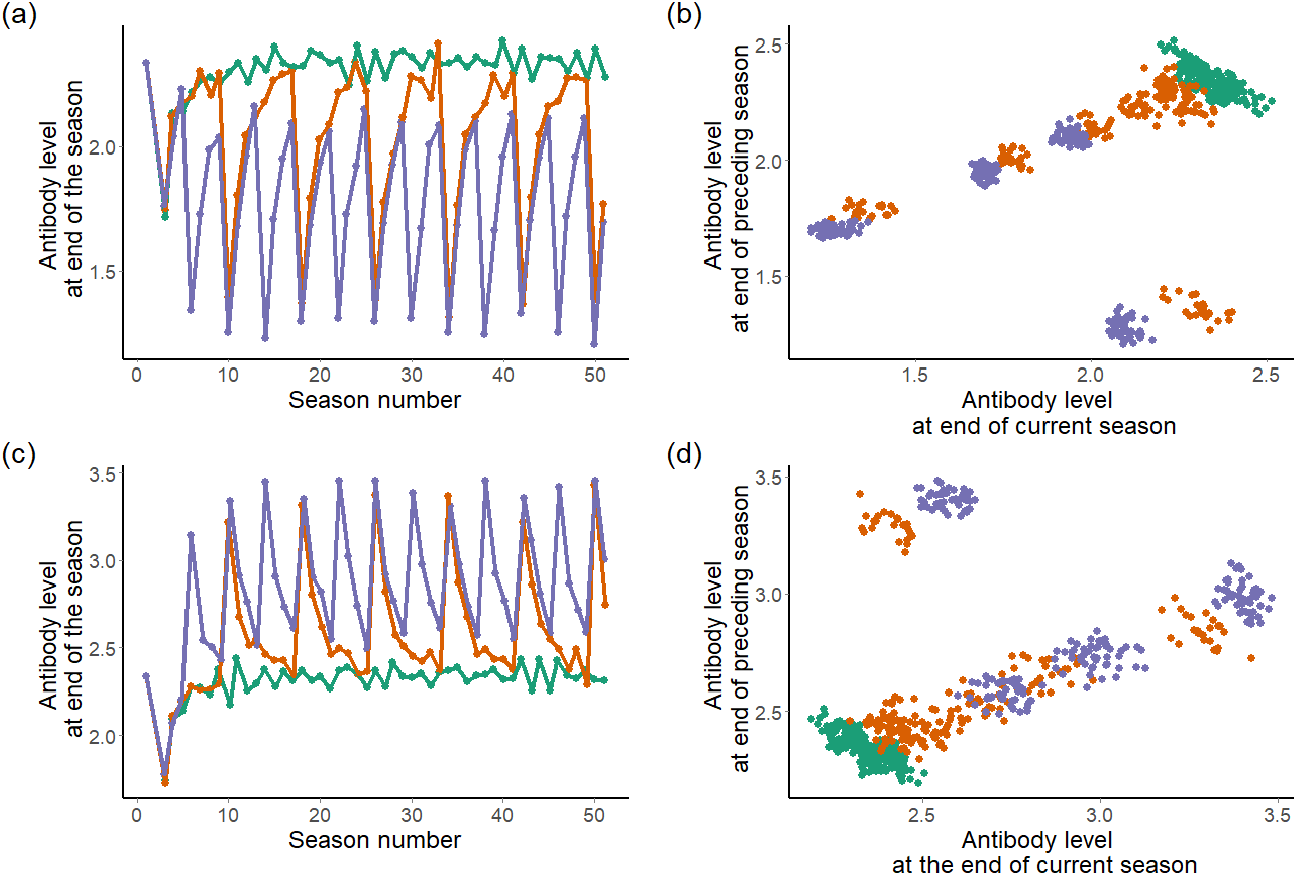
(a) End-of-season titer trajectories with an 8-year pattern of a missing season (orange), a 4-year pattern (purple) and with no missing seasons (green). (b) Change of titer from one season to the next. Displaced clusters represent the immediate effect of the missing boost. (c) End-of-season titer trajectories with an 8-year pattern of severe seasons (orange), a 4-year pattern (purple) and with no severe seasons (green). (d) Change of titer from one season to the next. Displaced clusters represent the immediate effect of the amplified boost.

### Lasting effects of a severe season

In a way analogous to the method in section above, we can model the after-effects of a severe season, by setting *β* = 2 for one season out of four, or one season out of eight, while *β* = 1 for all the remaining seasons. The results of this experiment mirror those of the ‘missing season’ experiment, but in the opposite direction. The orange lines in Figure 5(c) show how the titers in the 8-year pattern eventually settle down to rates comparable to the unmodified sequence, while the 4-year pattern shown by purple lines in Figure 5(c) retains elevated titers throughout. Figure 5(d) shows how the titers of the two severe-seasons trajectories gradually decrease towards the unmodified (no severe seasons) pattern.

Both experiments, one invoking a missing season and the other creating a severe season, suggest a gradual return to normal patterns over few years, rather than a disproportionate reaction in the immediately following season. The evidence in Figure 1 is not unequivocal, but corroborates this, at least partially.

## Discussion and conclusions

The core of our experimental model takes into account titer increases during a few months-long season of an infectious disease with fixed infectivity peak *μ*, plus a continuous process of waning, expressed by a negative exponential with parameter *ω*. These two processes can reproduce the cycle of boosting and waning characteristic of recurrent seasonal infectious disease. Iterating this model, however, is not suitable for generating long term trajectories of an epidemic. Mathematically, it is a contractive process that would quickly converge towards a sequence of identical seasons. Replacing the assumption of fixed peak times with a random choice among several months, however, modified the contractive tendency, so that simulations could explore the variation in titer as a function of the peak location parameter *μ*. This revealed a strong association between shift of peak month and change of titer from one season to the next, so that Figure 3(b) expresses a mathematically-based explanation of the negative correlated pattern discovered in national statistics (Figure 1(b) and (d)).

Introducing an element of causality into this association, we trained the model so that a lower titer in the previous year would lead to an early onset of the epidemic in the current year, via a slight bias in the random selection of the peak month, while a larger titer would delay the season. The pattern that emerged from this training then allowed us to establish a prediction rule [9] so that from the titer at the end of one season, we can predict the timing of the next one, in terms of a probability distribution of the timing of the peak month.

Inspired by predictions that the dramatic drop of infections by non-COVID-19 respiratory viruses like influenza and RSV in the 2020-2021 seasons would be followed by a strong resurgence in the following year, we adapted our model by setting the amplitude (severity) parameter *β* to zero for one year out of four or one year out of eight and observed how such perturbations affected subsequent years. These experiments showed that after each of the “missing” years, the trajectories of the post-season titers inevitably recovered towards the usual pattern, largely in the case of the four-year cycle, but completely in the case of the eight-year cycle.

To study the effects of a year with increased severity, we doubled the parameter *β* for one year out of four or one year out of eight and observed how such perturbations affected subsequent years, similar to the experiment with missing years. These experiments showed that after each of the severe years, the trajectories of the post-season titers inevitably reverted towards the usual pattern, largely in the case of the four-year cycle, but completely in the case of the eight-year cycle. This timing is of course dependent on our choice of parameters and protocols: *ω, β*, the training protocol, and predicting the distribution of *μ*. Nevertheless, it illustrates the kind of investigation possible with our titer-based modelling.

In further work, we could consider population heterogeneity by coupling several models representing antibody dynamics in different subgroups. In contrast to the movement of individuals in classical compartmental models, our model would not involve the transfer of titers between subpopulations. The effect of one compartment affecting another would modeled in the choice of *μ* and/or *β* in one compartment depending on the recent state of the process in a ‘neighbouring’ compartment, such as age group, spatial proximity, health workers versus general public or interacting occupational groups.

In this work, we have not assumed anything about viral strains. Apparent waning over several seasons may reflect mutational drift or selection in the antigen, rather than immunological processes per se. This does not distract from the pertinence of our model as a basis for analyzing the cyclical behaviour of boosting and waning. Many other factors may enter into the timing of the induction of an infectious season, such as climate, demographic changes, antigenic shift in the pathogen, changes in transmission patterns [9], and others. Whether or not post-season residual titer levels remain a likely driver of onset and peak times and severity remains an open question, on the basis of our work on influenza A.

## Data Availability

All data, code, and materials used in the analysis are available online at https://github.com/SiyuChenOxf/residualimmunity

https://github.com/SiyuChenOxf/residualimmunity

## Acknowledgement

We thank Yves Bourgault for key pointers leading to our discussion of dynamical systems. S.C. is funded by a Postdoctoral Research Fellowship in the High Meadows Environmental Institute, Princeton University. D.S. is funded by NSERC Discovery Grant RGPIN 5212-2022, the Canada Research Chair in Mathematical Genomics, and a University of Ottawa Distinguished University Professorship.

